# Acute Effects of Coherence Breathing on cardiopulmonary and autonomic responses: a randomized crossover study protocol

**DOI:** 10.64898/2026.06.30.26356965

**Authors:** Gisena Scotto Diclemente, Silvia Sole, Jamie Pigman, Tamara Rial-Faigenbaum

**Author notes:** **CORRESPONDING AUTHOR** Silvia Sole, PhD.

## Abstract

**Background:** Cardiopulmonary exercise testing (CPET) is a gold-standard test used to evaluate cardiopulmonary fitness and overall health by measuring physiological responses such as oxygen consumption during exercise. While traditional CPET warm-ups are typically low-intensity aerobic activities, alternative methods like coherence breathing may also prepare the body by influencing autonomic regulation. Breathing-based interventions have shown potential to improve heart rate recovery and performance, and heart rate variability (HRV) serves as a useful non-invasive indicator of autonomic nervous system activity. However, there is limited research on how brief breathing exercises before CPET affect outcomes. This study aims to investigate the effects of coherence breathing on oxygen uptake, HRV, and post-exercise heart rate recovery

**Objective:** This study will aim to compare the acute cardiopulmonary and autonomic responses of coherence breathing versus spontaneous breathing immediately preceding cardiopulmonary exercise testing (CPET) in recreationally active healthy adults.

**Methods:** This study will be a randomized counterbalanced crossover design. Healthy adults aged between 19 and 45 years of age will complete two separate CPETs over two non□consecutive test days (between 48 hours and 7□days). During each visit, participants will complete five minutes of slow-paced coherence breathing (6 breaths per minute) or spontaneous breathing at normal breathing rate, followed by an incremental treadmill CPET protocol up to maximal exertion. HRV will be assessed at baseline, during the breathing interventions, and during cool-down for 5 minutes using the Emwave Pro Plus software. Gas exchange during the CPET protocol will be measured continuously using the VO2 Master Pro system. immediately after, and after 5 minutes of resting. The primary outcomes will be peak oxygen consumption and heart rate variability indices. Secondary outcomes will include heart rate recovery, peak heart rate, time to exhaustion, rate of perceived exertion and readiness, blood pressure, tidal volume, peak ventilation, and respiration rate. Analyses will use linear mixed-effects models and paired comparisons.

**Discussion:** This protocol will determine whether pre-exercise coherence breathing can improve cardiopulmonary and autonomic nervous system responses to maximal performance. Findings may have practical implications for exercise testing and performance procedures as well as improving our understanding of pre-exercise breathing strategies for priming the autonomic and cardiopulmonary systems.

**Ethics/registration:** IRB SP2645 (May 15, 2026), clinical trials Identifier: NCT07650279

## BACKGROUND

Cardiopulmonary Exercise Training (CPET) is commonly used in clinical and research settings to assess health status and aerobic fitness. Physiological responses to submaximal and maximal aerobic exercise can provide important information about cardiopulmonary function and fitness level. In addition, CPET can assist in establishing baseline biometric data, designing an exercise program, tracking progress, and encouraging ongoing participation in physical activities (1,2). CPET is considered the gold standard because it combines standard graded exercise testing with simultaneous ventilatory respired gas analysis, which is commonly known as oxygen consumption (VO_2_). CPET also allows us to better understand the responses from a variety of exercise training programs (3,4,5). The American Heart Association proposed in 2016 that cardiopulmonary fitness should be considered as a clinical vital sign and routinely assessed in clinical practice, so CPET is demonstrated to be one of the most important tests when thinking of improving the general population’s health and non-communicable diseases’ prevention (6,7).

Warm-ups are used to increase body temperature, heart rate, blood flow, and respiration rate in the adult and young population (8,9). Traditional warm-ups for CPET are done in aerobic mode, just walking or cycling at low intensities, before an exercise test to evaluate cardiopulmonary status. However, other modes of training might also promote effective warm-up effects on the body before a CPET. Emerging evidence suggests that breathing-based interventions can acutely influence autonomic regulation and ventilatory control (10). Specifically, coherence breathing training has been found effective for heart rate recovery and subjective performance in athletes (11,12).

On the other hand, Heart Rate Variability (HRV) provides a non-invasive marker of autonomic nervous system activity and may offer additional insight into pre-exercise strategies and physiological state (13,14, 15). However, limited research has examined whether brief slow-paced breathing interventions before standardized exercise influence CPET outcomes (16). This study evaluates the potential role of slow-paced breathing using the coherence technique on oxygen uptake variables, HRV, and heart rate recovery.

## OBJECTIVES

The objective of this study is to examine whether a brief pre-exercise coherence breathing influences cardiopulmonary and autonomic responses during CPET in healthy, recreationally active young adults. The aims are as follows: i) to compare physiological responses following a seated coherence breathing intervention versus quiet spontaneous seated rest before standard treadmill CPET; ii) to analyze primary outcomes such as oxygen uptake kinetics and HRV; iii) to analyze secondary cardiopulmonary responses during exercise, time to exhaustion, peak heart rate, and heart rate recovery, blood pressure, or perceived exertion. iv) to improve the understanding of the influence of autonomic priming strategies on exercise outcomes.

## METHODS

### Study design

This study is a randomized, counterbalanced two□condition crossover employing a Latin□square order. Two visits are scheduled ≥48□h and ≤7□d apart, at the same time of day under controlled environmental conditions. Estimation of sample size was done with an a priori power analysis run by the software G-Power (version 3.1.9, Heinrich-Heine-University, Düsseldorf, Germany), with a desired power level of 0.90, an alpha level of 0.05, and an effect size of (Cohen’s *d* = 0.80) using a two-tailed paired samples t-test to compare mean differences between two conditions (17). A total sample size of 19 participants was required.

### Participants

Inclusion criteria for participants include: a) between 19 to 45 years of age; b) accumulate a minimum of 3 hours per week of moderate to vigorous physical activity measured by the IPAQ questionnaire (18); c) be able to participate in moderate to vigorous physical activity as determined by the Physical Activity Readiness Questionnaire, PARQ (19). Exclusion criteria are as follows: a) accumulate less than 3 hours per week of moderate or vigorous physical activity; b) have a health condition that is a contraindication for performing maximal CPET including cardiovascular disease, pulmonary issues, metabolic conditions, neuromuscular disorders, orthopedic limitations, or acute illness that impedes performing CPET; d) not completing the study procedures or not providing informed consent and; e) not be able to participate in moderate to vigorous physical activity as determined by the PARQ-2023 questionnaire.

Before the start of the study, all participants will be informed about the study’s aims and procedures and will sign the informed consent form. This study was approved by the Institutional Review Board at Monmouth University. (SP2645, May 15, 2026). All participants will sign a consent form verifying that they meet the inclusion criteria and understand the benefits and risks of the study. All collected data will be deidentified. Data will be sourced and gathered respectfully and inclusively, adhering strictly to the inclusion and exclusion criteria listed above and following ethical principles outlined in the Declaration of Helsinki.

### Experimental Design

Participants will report to the Human Performance Laboratory of Monmouth University from the Department of Health and Physical Education situated at the Monmouth University Graduate Center, room 222 (West Long Branch, NJ). They will come on two nonconsecutive days (within 1 week maximum) at the same time of day and under the same controlled temperature and humidity conditions. Participants will be instructed not to participate in vigorous exercise within the 48 hours before the testing and to refrain from caffeine and alcohol consumption before the day of testing, as well as to avoid eating for 3 hours before the testing and to use the same shoes for both trial sessions.

A randomized, counterbalanced crossover design will be employed. Participants will complete both experimental conditions (coherence breathing and spontaneous breathing), with the order of conditions determined by computer-generated randomization (www.randomizer.org). Participants will be given standardized instructions before the start of the test and the same verbal encouragement during both testing procedures. The procedures are explained in figure 1

**Figure 1.**
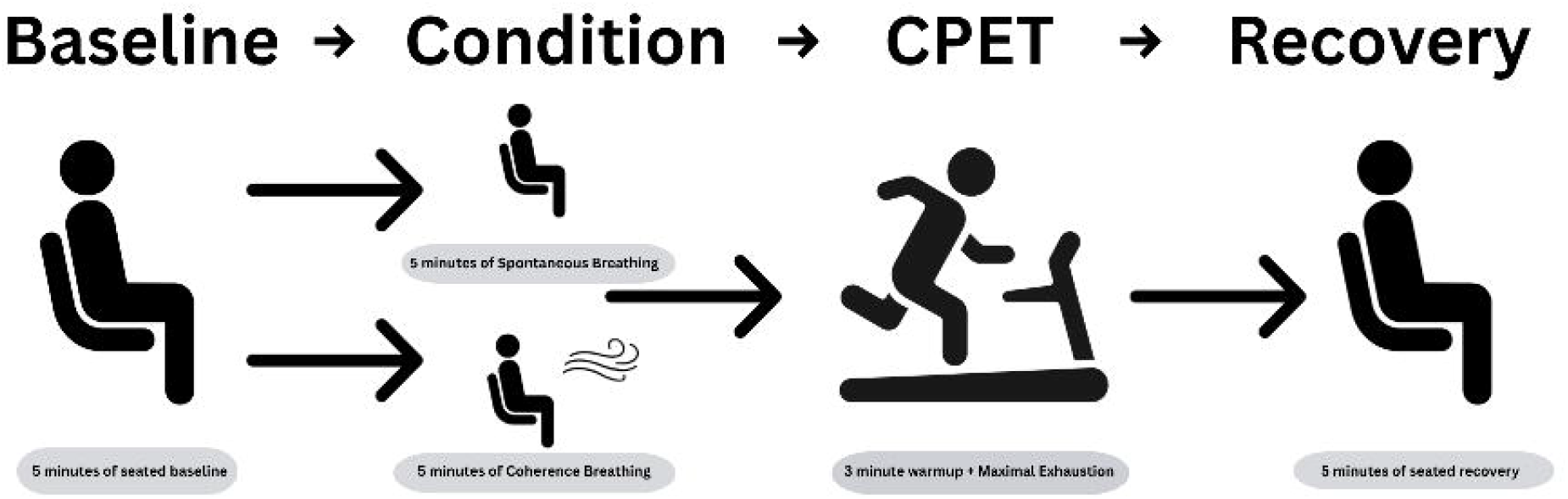

#### Coherence Breathing protocol

Participants will complete a 5-minute seated coherence breathing before CPET. This condition will be paced at a slow rate at approximately 6 breaths per minute (5 seconds per inhalation and 5 seconds per exhalation). Participants will follow a visual breathing pacer displayed on a TV monitor to maintain the prescribed breathing rhythm by using the Free Breath Pacer Resonance Frequency Breathing Tool (www.allos.one). Participants will be instructed to follow the breathing rate according to the visual stimuli of an expanding and contracting circle while maintaining a relaxed seated position. HRV will be continuously monitored through both breathing interventions.

#### Spontaneous Breathing Protocol

Participants will be seated comfortably in a chair for 5 minutes before CPET. During this condition, participants will be instructed to breathe normally at their own pace without attempting to control the depth or rate of breathing.

#### Cardiopulmonary Exercise Testing

The treadmill protocol test to be used is adapted from the protocol proposed by Green et al. (20), which is a valid and reproducible exercise protocol designed for CPET and previously assessed in healthy adult populations (20). Consists of 9 stages with gradual increments in speed and incline. The test consists of progressively increasing treadmill speeds and inclines over two-minute stages until voluntary exhaustion or when maximal heart rate has been achieved. The initial stage begins at a low workload, and each subsequent stage increases the speed (mph) and incline (%). HR and RPE will be measured at each stage to confirm equal physiologic and perceived workload during conditions. After the conclusion of the CPET, the participant will have a cool-down by sitting on a chair for 5 minutes to recover and measure baseline physiological variables (HR, HRV, BP, RPE).

Exercise intensity will be determined by using the Gellish et al. (21) predicted HR maximum (HRmax) formula: HRmax = 207− (0.7 x age). This formula was selected because it is more accurate in the prediction of HRmax in well-trained individuals (21, 22). A participant’s peak VO□ will be reached when one of the following conditions described by ACSM’s guidelines in exercise testing (23) is met: a) an RPE between 9 and 10; b) HRmax reached as determined by the predicted Gellish et al. (21) formula; c) final respiratory exchange ratio of 1.1; d) steady state of VO□ despite increasing intensity. We will measure the total time of exhaustion of the test and the last stage of CPET completion.

#### Outcome Measures

All anthropometric variables will be measured following standardized procedures as described by the American College of Sports Medicine (23). Height will be measured using a wall-mounted stadiometer, and body composition will be measured using the body composition analyzer InBody 380. Blood pressure will be assessed by an automated sphygmomanometer. The RPE will be assessed using the Borg Scale (1-10) (24). RPE will be taken at the beginning of the session in a seated position as baseline measurements of resting values. After the breathing conditions, immediately after the CPET protocol, and after 5 min of rest, these vital values will be retaken.

HRV will be taken with Emwave pro plus (www.heartmath.org) which is a computer software that collects HRV data through a pulse sensor placed on the participant’s earlobe. Participants will be instructed to remain seated for 5 minutes to collect baseline HRV data and to refrain from making any rapid body movements. The same instructions and procedures will be given for HRV assessment during the breathing conditions (spontaneous vs coherence) and during the cool-down phase immediately after the CPET. The EmWave Pro Plus enables the import of raw RR intervals into Microsoft Excel for further statistical analysis.

A calibrated metabolic system (VO□ Master Pro Systems, Vernon, Canada) will be used to assess gas exchange. The VO□ Master Pro is a portable device that operates through breathCbyCbreath gas exchange analysis. This analyzer is composed of a differential pressure flow sensor, an OC analyzer (galvanic fuel cell sensor), which measures VC_E_ (large: 40 to 220□L·min−1, medium: 30 to 160□L·min−1, and resting: 5 to 40□L·min−1). The VO_2_ Master Pro system will be calibrated before use with a one□point calibration that uses room air and a 3□L syringe to calibrate the O□ and flow sensors. All metabolic variables will be collected via Bluetooth to an iPad tablet (Apple, Inc., Cupertino, CA, USA) equipped with the VO□Master Pro mobile app (Vernon, Canada, vo2master.com) for storage and later download of data. A Polar heart rate monitor/strap (Polar Electro Oy, Kempele, Finland) is integrated into the V0_2_ Master unit to record HR continually. The portable Bluetooth system allows users to perform suspension dynamic exercises without being impaired by cables. Participants will wear the VO□ Master Pro system during both trial conditions. The VO_2_ Master has previously shown acceptable validity and test-reliability for ventilatory assessment (25, 26).

### Statistical Methods

VO_2_ Master Pro data will be reintegrated to 30-second intervals for analysis and exported to Microsoft Excel. The raw RR intervals from the Emwave Pro Plus will be exported into Microsoft Excel. Descriptive statistics (mean□±□SD) will be reported. Primary outcomes will be analyzed using a linear mixed-effects model (LMM) with Order (day 1 vs day 2) as fixed effects and participant as a random intercept. Normality of paired differences will be assessed with the Shapiro-Wilk test; if normality is violated, a Wilcoxon signed□rank test will be used. A paired t-test will be used to compare the study cardiopulmonary and HRV variables between the two breathing conditions. Effect sizes will be reported as Cohen’s d (27) for parametric tests or matched-pairs rank biserial correlation for non-parametric tests, with 95% confidence intervals. Analyses will be conducted in Jamovi (v2.3.21, Sydney, Australia) with the GaMLj (v2.8.0) module, and HRV data will be analyzed using the Kubios software. Statistical significance will be set at *p=* 0.05.

## Data Availability

All data produced in the present study will be available upon reasonable request to the authors

## DECLARATIONS

### Ethics approval and consent to participate

This study was approved by the Institutional Review Board of Monmouth University, New Jersey, US (May 15, 2026; Protocol SP2645).

### Consent for publication

Not required.

### Availability of data and materials

Not applicable at this stage.

### Authors’ contributions

TR is the guarantor. Conceptualization: TR Methodology: TR, SS. Writing—original draft: TR, SS. Review & editing: TR, RF, JP, GSC. All authors have read and approved the final protocol.

### Trial registration

This study was registered at ClinicalTrials.gov (Identifier: NCT07650279).

## Acknowledgements

The Monmouth University Summer Scholars Program and mobility Grant supported this research

